## Supplementary material for "The psychosocial impact on frontline health and social care professionals in the UK during the COVID-19 pandemic: a qualitative interview study": Figure 2

Communication challenges

Virtual consulting

Difficult conversations

Support structures

Team unity

Leadership

Social support

Work-related stressors

Need to protect loved ones

Public not following rules

Increased workload and changing work conditions

Uncertainty of risk to patients

Resilience

Proactive coping

Accepting uncertainty

Increased sense of purpose and reward

Personal growth

Slowing down

Increased reflection

Improved non-work relationships

Figure 2: Themes and sub-themes
