## Supplementary material for "The psychosocial impact on frontline health and social care professionals in the UK during the COVID-19 pandemic: a qualitative interview study": Table 1

Table 1: Characteristics of the health and social care professionals

| Number of participants | 25 |
| --- | --- |
| Profession | Hospital doctor (6)  GP (4)  Hospital nurse (3)  Social worker (3)  Home carer (2)  Care home carer (2)  Assistant psychologist (1)  Community mental health nurse (1)  Practice nurse (1)  Counsellor & psychotherapist (1)  Physiotherapist (1) |
| Age | Range 26-65 (Mean 39) |
| Gender | Male 5  Female 20 |
| Ethnicity | White British 17  Asian 3  Black British 2  White & Asian 1  White Irish 1  White Other 1 |
