## Supplementary material for "The psychosocial impact on frontline health and social care professionals in the UK during the COVID-19 pandemic: a qualitative interview study": Figure 1

- How would you describe your social life now that social distancing measures have been brought in because of Covid-19?
- In what ways has your work life been impacted by the Covid-19 pandemic?
- How do you feel about the changes that have been brought about by Covid-19? Have they had any impact on your mental health or wellbeing?
- Have there been any positive experiences for you resulting from the Covid-19 pandemic?

Figure 1: Examples of Questions in the Topic Guide
