## Supplementary material for "The psychosocial impact on frontline health and social care professionals in the UK during the COVID-19 pandemic: a qualitative interview study": Interview guide

**Draft topic guide: Health and Care Workers**

**Ask to describe ‘normal life’ – before the crisis, and now**

- Employed? Type of job, hours etc,
- Full time parent or carer?
- Who you normally live with, does this change, separated/ extended family?
- Whether you would usually have done any type(s) of regular exercise (whatever they perceive as exercise including walking/gardening)

**SOCIAL LIFE**

**What was your social life before the Covid-19 pandemic? Has this changed? If so, what has been the impact of Covid-19 on your social life?**

- How would you describe your social network before Covid – for example size, types of people, types of relationships, do they live with you, nearby or further away, how often do you see each other, how well do you know each other? How do you interact, face to face, online or social media? Describe some of your common socialising activities. **Has this changed? What has the impact of Covid been on your social network?**
- Can you tell us about any ways your social networks/ friendship groups influence you, such as peer pressure, or encouraging you to get involved in things? Do you compare your life to theirs?
- Could you describe any community participation or volunteering participation before Covid? **Has this changed? If so, what has been the impact of Covid-19 on community participation/volunteering participation?**
- Could you describe the social support you have before Covid? (such as emotional support, advice and information, someone to help you with money or milk/bread/essentials) **Has this changed? If so, what has been the impact of Covid-19 on your social support?**
- **Social engagement (social roles, bonding, attachment) (pre- and post- Covid)**

**WORK LIFE**

**How would you describe your work life before the Covid-19 pandemic?**

Prompts include:

- Describe a typical day?
- Describe your work environment prior to the crisis
- How much autonomy did you have in your role?
- Did you find your job rewarding?
- Did you feel able to do your job to a high standard?
- Did you enjoy your job?
- Describe your sense, if any, of team unity or disunity prior to this crisis?
- How able were you to follow organisational rules and how did you feel about this?
- Normally did you feel safe at work? In what way?

**How would you describe your work life since the Covid-19 pandemic? Please tell us about this**

- Describe a typical day now – how have common work practices changed? Have you adapted your work in response to Covid-19 (e.g. delivery, operating hours, change of products/production methods)
- Describe your overall work environment now
- How much autonomy do you feel you have at the moment and how has this changed?
- Are you finding work rewarding at the moment?
- Do you feel able to do your job to a high standard – has this changed since the crisis?
- Enjoyment – do you currently enjoy your job?
- Describe your sense, if any, of team unity or disunity during this crisis?
- How able are you to follow organisational rules and how do you feel about this?
- Do you feel safe? If this has changed, how?

**MENTAL HEALTH**

**How do you feel about the changes that have been brought about by Covid-19?**

**Have they had any impact on your mental health or wellbeing? Please tell us about these**

- What are the things most bothering you at the moment (work or outside of work)?
- What have been the major triggers/causes of any mental health or wellbeing issues?
- How have government guidelines or organisational guidelines impacted your mental health or wellbeing?
- Have you experienced any impact on positive emotions? (prompts: how deeply you can engage with what you are doing, sense of meaning/ purpose, relationships with others, how well you are managing and feelings of control over your situation?)
- Has there been any impact on your sense of identity?
- Have you experienced any negative psychological feelings? (prompts: such as shame, guilt, lack of pleasure, anxiety, worry)
- Please tell us about any physical symptoms due to being stressed or anxious? (prompts: fatigue, sleep problems, pain, illness symptoms, palpitations)

**Have you been doing/ planning anything to help with this?**

- How has your support been, from friends/family? From work colleagues/your organisation?
- Connecting with family or friends online
- Online groups?
- Hobbies/ Reading
- Exercise at home <ask about what they have been doing and if there are specific resources they have found useful to exercise>
- Volunteering
- Other engagement

**Why are you doing/ not doing these things?**

- Helpful/ not helpful – please tell us why
- Enjoyable
- Good for mental health/ wellbeing
- Can’t get online, not connected, not comfortable, affordability, confidence in using/ skills
- Skills in using the internet/ communication software
- Living arrangements/ Work/ caring demands
- Peer support/ pressure
- Difficulties/ restriction in physical environment

**PROSPECTION**

**Has the pandemic meant that you have any worries for the future?**

- Worries about work/the future of your work?
- Worries for yourself? Anything not directly connected to work?

**How are these different from the worries you had before?**

- Sense of control/ powerlessness
- Severity of worries / perspective

**Will this change the way you live your life in future?**

- The way you connect with others
- How you look after yourself
- How you support others
- How you exercise?

Do you think there will be any changes to the way you work in the future? Why/why not?

Has this changed any of your priorities for the future?
